# The enigma of persistent left-handedness in humans: A potential solution

**DOI:** 10.64898/2026.05.20.26353697

**Authors:** Ruben C. Gur, Zhiqiang Sha, Tyler M. Moore, Monica E. Calkins, David R. Roalf, Kosha Ruparel, J. Cobb Scott, Anna Watters, Lauren J. Harris, Aaron F. Alexander-Bloch, Raquel E. Gur

**Affiliations:** Department of Psychiatry, Neurodevelopment and Psychosis Section, University of Pennsylvania Perelman School of Medicine, Philadelphia, Pennsylvania, USA; The Lifespan Brain Institute (LiBI) of Penn and Children’s Hospital of Philadelphia (CHOP); Department of Psychology, University of South Carolina, McCausland College of Arts and Sciences, Columbia, South Carolina, USA; Child and Family Health, Central Coast Local Health District, NSW Health, Australia; Department of Psychology, Michigan State University, East Lansing, Michigan, USA

## Abstract

The persistence of a left-handed minority of slightly over 10% of the population is enigmatic because it is associated with stigma, increased psychopathology, and cognitive deficits. In a community sample of 9,352 individuals (age range 8-21 years) with neurobehavioral assessments, left-handers (N=1,281, 673 male) indeed showed greater psychopathology and performed more poorly than right-handers (N=8,076, 3,839 male) on tests of executive function, memory, complex cognition, and social cognition, while excelling in motor speed. Furthermore, the variance was higher and within-individual variability (WIV) - the extent to which scores in the different domains varied within individuals - was higher in left-handers. Since low WIV indicates even distribution of abilities while high WIV reflects specialization in circumscribed areas, the finding indicates that left-handers are “neurocognitive specialists”. This combination of behavioral traits could confer resilience against natural selection pressures and help explain preponderance of left-handers in highly specialized professions requiring specific talents. Our findings encourage more research on left-handers, who are currently excluded from multiple brain behavior studies.

## Introduction

Most animals exhibit bilateral symmetry (order Bilateria), to match interactions with the environment where events occur and require response equally often on both sides (Corballis, 2010). Handedness - the behavioral preference to use one hand over the other - represents a departure from this symmetry. Though not exclusively hominoid(Hecht et al., 2025; Hopkins, 2018), handedness is among the most reliable behavioral traits associated with individual differences in human behavior (Ocklenburg, 2025). While most people (>87%) prefer using their right hand, around 11% of females and 13% of males are left-handed (Papadatou-Pastou et al., 2020; Perelle and Ehrman, 1994) (Gilbert and Wysocki, 1992; Peters et al., 2006; Vuoksimaa et al., 2009), a variant that has persisted since the middle Pleistocene epoch (ca. 425 000–180 000 YBP) (Corballis 2003, 2010).

The sacrifice of symmetric abilities has been explained by the parallel development of hemispheric specialization, whereby the left hemisphere, which controls the right hand, is structurally and functionally configured to serve language and complex reasoning. However, the evolutionary persistence of left-handedness presents a puzzle: throughout recorded history and across cultures, left-handedness has been stigmatized (Gofman 1963, Harris 1990, Masud & Ajmal 2012), and there is mounting evidence that it is associated with increased psychopathology (Packheiser et al., 2025) and cognitive deficits (Abbondanza et al., 2023; Grimshaw and Wilson, 2013). Why, then, haven’t left-handers disappeared entirely through natural selection? The answer may lie in the behavioral profile of left-handers, which could confer sufficient advantages in specific domains or in the configuration of their abilities. Satz and colleagues (Satz et al., 1985) hypothesized that the reason for the preponderance of left-handers’ pathology is base-rate statistical. If 90% of the population is biologically right-handed and 10% left-handed, and brain damage related to perinatal birth complications switches the laterality in 10% of the population, the result will be that about half of left-handers compared to only 5.6% of right-handers will have pathology-related reversed laterality. This model also explains the higher incidence of left-handedness in males than females (Raymond and Pontier, 2004), as brain-related effects of birth complications more often affect males (DiPietro and Voegtline, 2017). Furthermore, there is evidence for higher diversity in cognitive abilities among left-handers (Prichard et al., 2013; Searleman et al., 1984), suggesting greater variance of performance, including areas where left-handers outperform right-handers (Saari and Vuoksimaa, 2023; Sala et al., 2017, Peterson & Lansky, 1974), and some evidence suggesting greater modality-dependent within-individual variability in left-handers (Heled et al., 2025). However, the conundrum of the persistence of left-handedness remains unresolved, especially the contribution of unique neurobehavioral characteristics that may impact survival.

Unfortunately, most brain behavior studies exclude non-right-handers and hence left-handers are disproportionately underrepresented in meta-analyses (e.g. Bailey et al., 2020; Cano et al., 2025; Huang et al., 2025; Sriutaisuk and Franz, 2025; Willems et al., 2014). Identifying clinical and performance profile differences between left-handers and right-handers requires large community samples, with recruitment unbiased by handedness criteria and with comprehensive behavioral assessments. The Philadelphia Neurodevelopmental Cohort (PNC) offers such an opportunity, with neurocognitive data on over 9,000 individuals ages 8 to 21 years recruited and enrolled between 2009 and 2011.

Previous studies with this cohort established the reliability and validity of the clinical assessment and the neurocognitive testing tools: GOASSESS (Calkins et al., 2015) and the computerized neurocognitive battery – CNB (Gur et al., 2010, 2012, 2015, Moore et al., 2015; Roalf et al., 2014b). They revealed age effects and sex differences in speed and accuracy and their developmental trajectories (Roalf et al., 2014a, Gur et al., 2012, 2014). They also showed such effects on within-individual variability (WIV), defined as the variance among standardized scores within individuals (Roalf et al., 2014a). However, the PNC data include information on handedness yet the effects of handedness on psychopathology and neurocognitive performance have not been examined in this cohort. The PNC dataset uniquely enables testing the hypotheses that: 1} Left-handers show greater psychopathology across domains. 2(Left-handers show lower performance across domains. 3(Left-handers have higher variance of performance scores. 4) Left-handers have higher WIV. 5) These effects are systematically modulated by age and sex.

## Results

The sample with complete psychopathology and neurocognitive data consisted of 9,357 individuals (4,512 male (48.2%), 4,845 female (51.8%)), of whom 8,076 (86.3%) classified themselves at testing as right-handed (3,839 male (47.5%), 4,237 female (52.5%), and 1,281 (13.7%) as non-right handers (left or ambidextrous), henceforth referred to as left-handed (673 male (52.5%), 608 female (47.5%)). Expectedly, the proportion of left-handers was higher in males (14.9%) than in females (12.6%), X^2^=11.13, df=1, p<0.001.

To test Hypothesis 1 of greater psychopathology in left-handers we examined severity of reported psychiatric symptoms obtained by the standardized clinical interview (GOASSESS) in the domains of mood (anxiety and depression), phobias (including agoraphobia and specific phobias), externalizing behavior (including attention deficit hyperactivity, oppositional-defiant, and conduct problems), and psychosis. A Mixed-Model Repeated Measures (MMRM) analysis indicated a main effect of handedness group, F=9.33, df=1,9334, p=0.002, partial η^2^=0.0010, without effects of Domain or Group x Domain interaction. As can be seen in Figure 1, left-handers have higher severity across domains (Figure 1A), and this effect is seen in both males and females (Figure 1B). While the handedness x psychopathology domain interaction was not significant, it is noteworthy that the increased severity for left-handers was significant for externalizing behavior (p=0.003; Cohen d = 0.100) and psychosis (p=0.023; Cohen d = 0.094), but not for the other domains.

**Figure 1.**
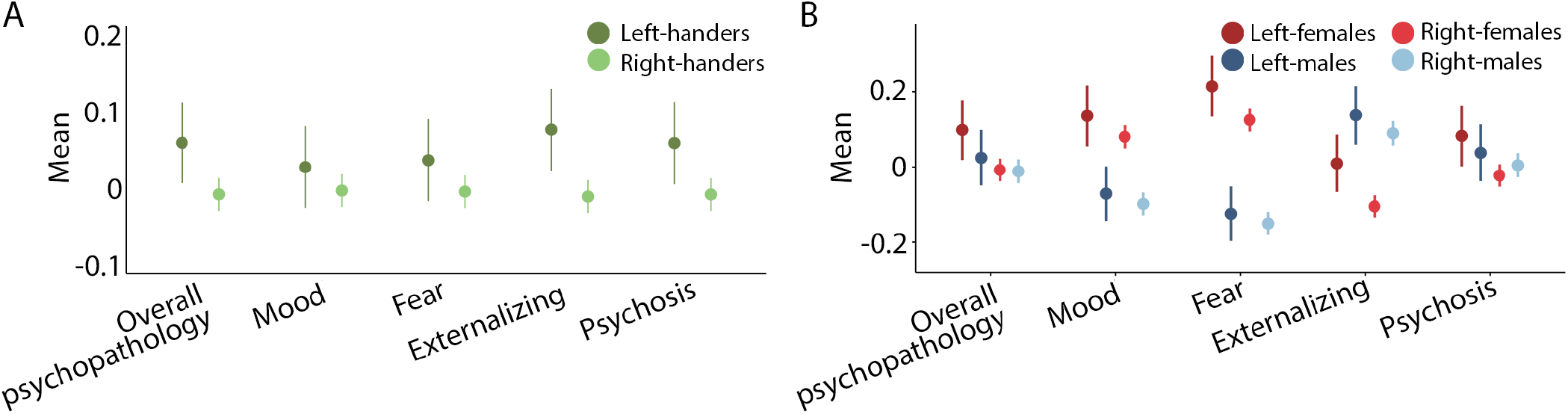
Symptom severity scores for the handedness groups: A. Comparing the two groups in overall symptom severity and in the domains of mood-anxiety, phobias, externalizing and psychosis. B. The groups separated by sex.

We next tested Hypothesis 2 of lower performance in left-handers using the accuracy and speed scores from the CNB. We used scores standardized across the entire sample (z-scores, with those for response time multiplied by -1 to make higher values in all scores reflect better performance), and for testing the hypothesis across the age range we regressed out age, age^2^, and age^3^. A Mixed-Model Repeated Measures (MMRM) analysis on the efficiency scores indicated a main effect of handedness group, F=35.90, df=1,9091, p<0.001, partial η^2^=0.0039, with lower overall efficiency in left-handers, and a handedness x test interaction, F=12.66, df=13,118724, p<0.001, partial η^2^=0.0014, indicating that this difference was not equally evident across tests. As can be seen in Figure 2, left-handers performed less efficiently across domains except for attention and motor speed, where they outperformed their right-handed counterparts, and working memory, spatial memory, and spatial cognition, where the effects were small (Figure 2A). The effects were present in both males and females (Figure 2B).

**Figure 2.**
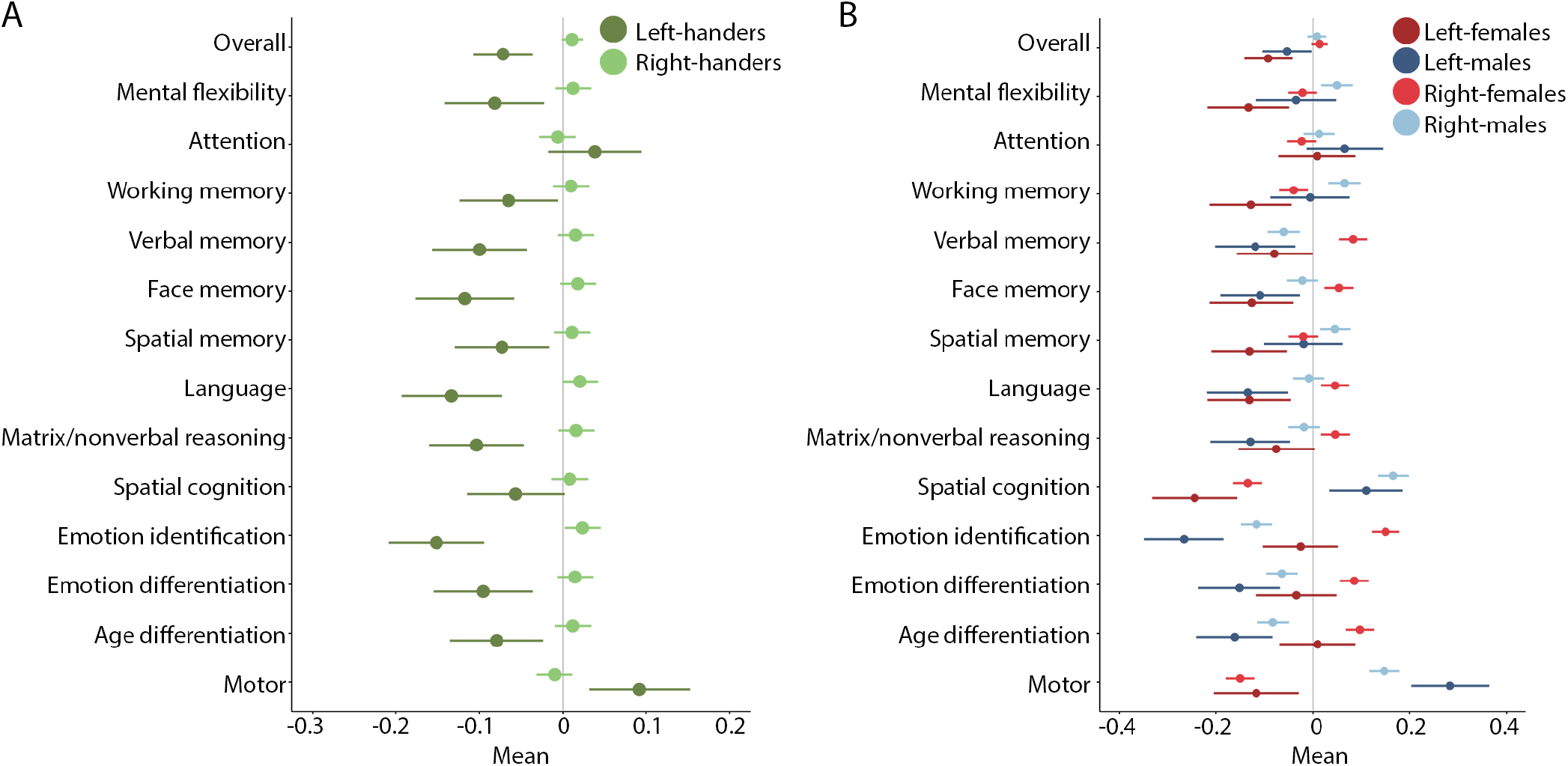
Neurocognitive profile for the handedness groups: A. Comparing the two groups on overall efficiency and in the domains of executive functions (Abstraction and mental flexibility, sustained attention, working memory), episodic memory (verbal, facial, spatial), complex cognition (language mediated logical reasoning, nonverbal abstract reasoning, spatial processing), social cognition (emotion identification, motion intensity differentiation, age differentiation), and praxis (sensorimotor and motor speed). B. The groups separated by sex.

Follow-up analyses adding accuracy vs. speed as a vector revealed an interaction of handedness x test x accuracy vs. speed, F=7.10, df=11,228433, p<0.001, partial η^2^=0.0003, indicating that the effects of handedness were stronger for accuracy for some tests and for speed in others. As can be seen in Figure 3A, left-handers showed deficits for speed and not accuracy for the verbal memory and emotion identification tests, while their deficit was specific for accuracy and not speed for the nonverbal reasoning test and they outperformed right-handers on attention speed. There was also a handedness x sex x accuracy vs. speed interaction, F=12.28, df=1,228480, p<0.001, partial η^2^=0.0001, indicating that the effects of handedness were stronger for accuracy in females than in males and about equal for speed in the two sexes (Figure 3B).

**Figure 3.**
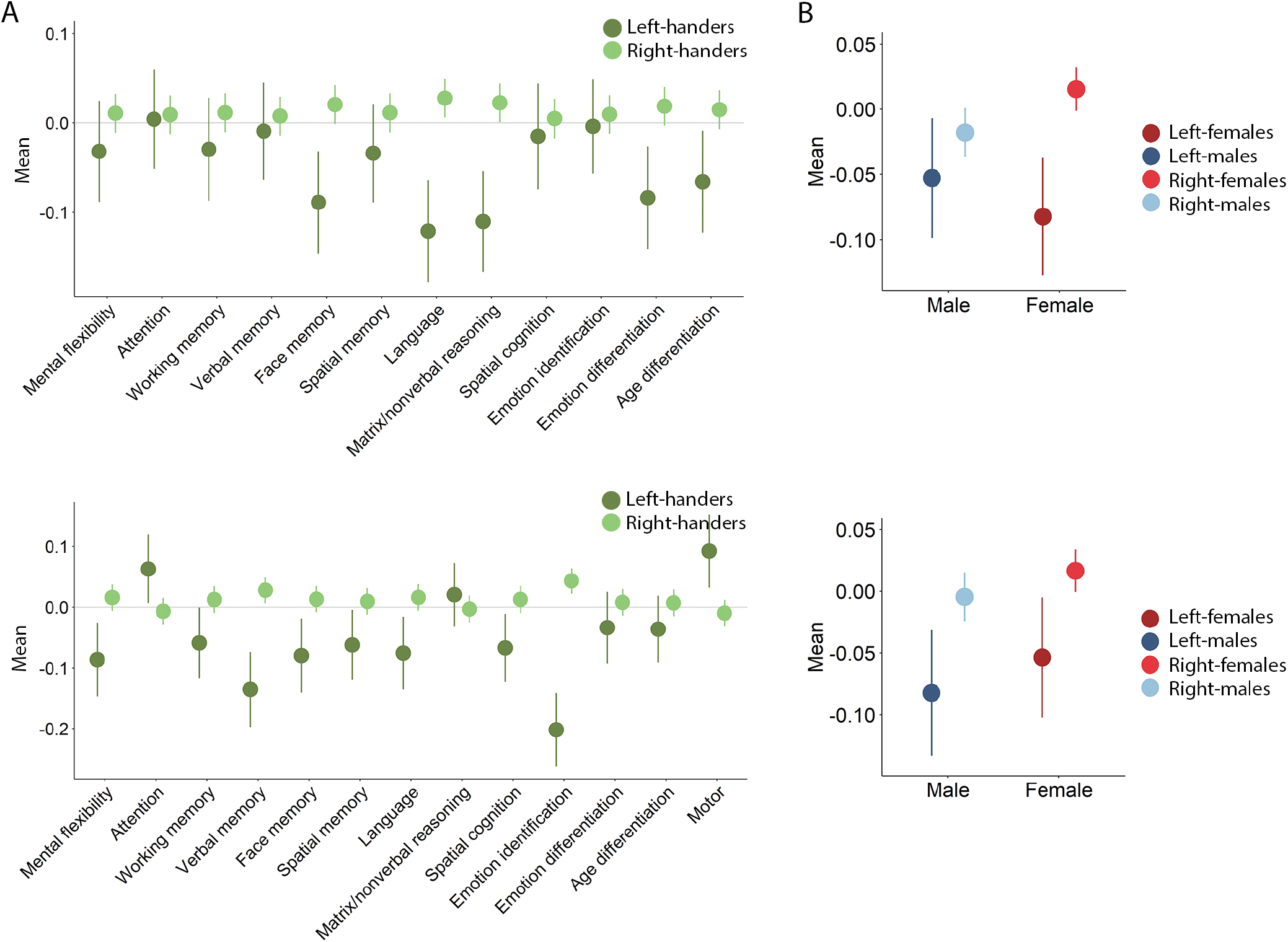
A. Neurocognitive profile for the handedness groups by accuracy (top panel) and speed (bottom panel) illustrating the handedness x test x accuracy vs. speed interaction. B. Cognitive test scores by accuracy (top panel) and speed (bottom panel) for males and females illustrating the handedness x sex x accuracy vs. speed interaction.

Hypothesis 3 of higher variance in the left-handed group was tested with the levene test and was supported, with left-handers showing higher variance F=1.10, p=0.029. The handedness groups distributions did not differ in kurtosis, skew, or bimodality (Hartigan dip test).

Hypothesis 4 of handedness differences in WIV was tested by taking the standard deviation of all accuracy and speed scores. The hypothesis was supported, with higher WIV for left handers, F=14.88, df=1,9232, p<0.001, partial η^2^=0.0016. There was also a handedness x accuracy vs. speed interaction, F=4.78, df=1,64349, p=0.029, partial η^2^=0.0001, indicating that the effects of handedness on WIV were more pronounced on speed than accuracy.

We next examined Hypothesis 5 by charting the developmental trajectories of the handedness differences as modulated by sex. To examine the developmental trajectories of clinical symptoms and performance we used the z-scores standardized on the entire sample and divided it by 2-year age groups (range of 8 to 21 years). For the clinical symptoms, adding Age group and Sex as grouping factors to the MMRM, we found significant effects of handedness, left-handedness associated with more severe psychopathology, F=9.33, df=1,9334, p=0.002, partial η^2^=0.0010, age group effect, F=123.07, df=6,9334, p<0.001, partial η^2^=0.0733, age associated with increased severity in all symptom domains, and no higher-order interactions.

As can be seen in Figure 4, Mood and Anxiety symptoms, as well as specific phobias, were more prominent in females than males across the age range, yet they were more prominent in left handers in both sexes, especially in late adolescence. Externalizing symptoms were more prominent in males than in females across the age ranges, but again both male and female left-handers showed greater severity, especially during adolescence (although severity dropped in left-handed females toward early adulthood). For psychosis, right-handers showed the expectedly higher severity in males across the age range but especially toward young adulthood. Left-handers, both male and female, show increased psychosis symptoms especially during adolescence with further increase toward adulthood.

**Figure 4.**
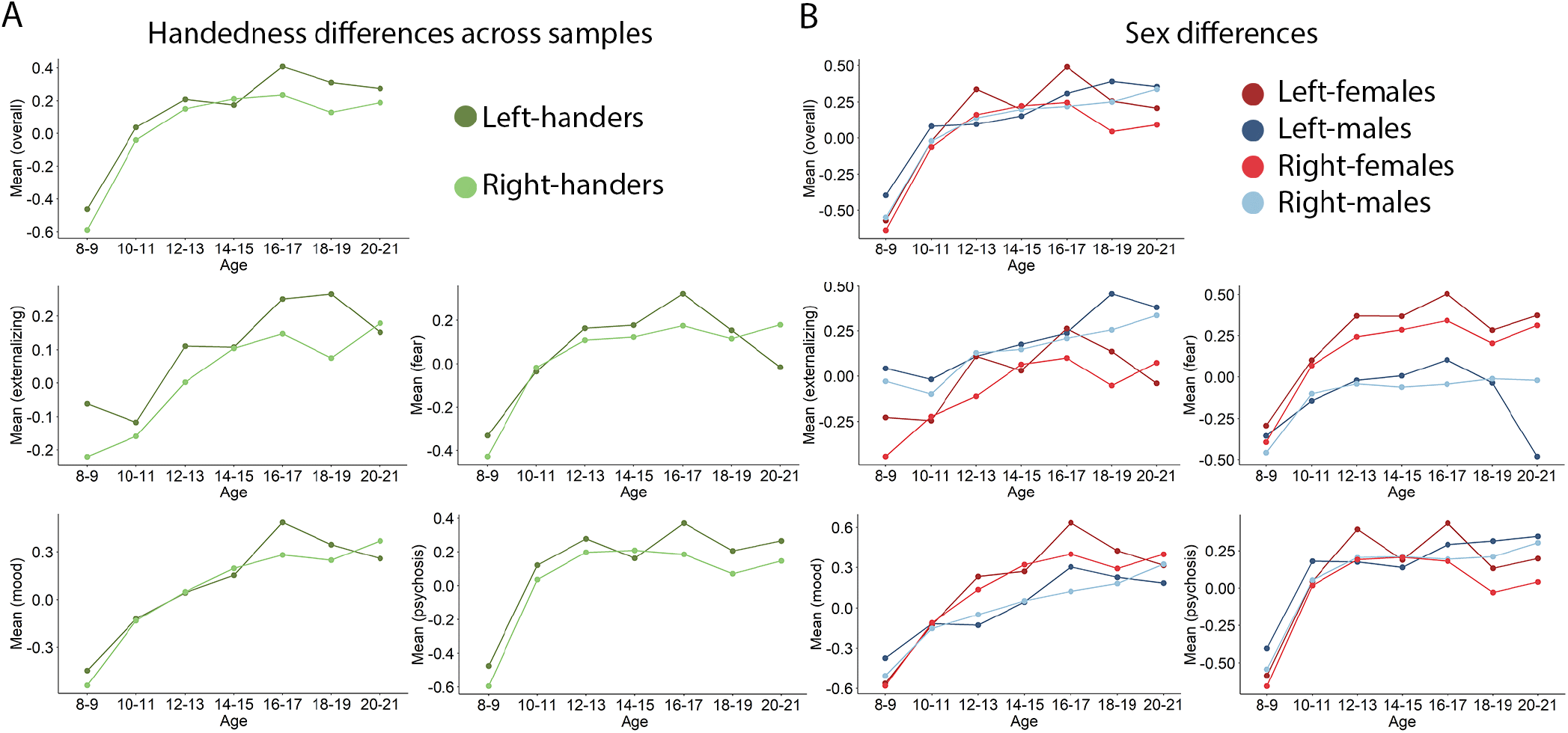
Age trajectory for psychiatric symptoms associated with handedness: A. Comparing the two groups in the domains of externalizing, mood-anxiety, phobias, and psychosis across the age range of 8 to 21 years. B. The groups separated by sex.

We next examined the developmental trajectories of cognitive performance in the two handedness groups. The MMRM showed significant effects of age group, older age associated with improved performance efficiency, F=728.18, df=6,9293, p<0.001, partial η^2^=0.3198, and one handedness x age group x accuracy vs speed interaction, F= 7.39, df=6,228513, p<0.001, partial η^2^=0.0002. As can be seen in Figure 5A, both groups showed the expected improvement with age for both accuracy (top panels) and speed (bottom panels). However, left-handers performed more poorly for speed, especially at the earlier ages. While the 4-way interaction of handedness x sex x age group x accuracy vs. speed was only marginal (F=1.84, df=6,228496, p=0.086), it appears that accuracy and speed showed somewhat different trajectories for left-handed and right-handed males and females. For accuracy, while female left-handers showed lower performance across the age span, male left-handers caught up with their right-handed counterparts by young adulthood. For speed, female left-handers caught up with their right-handed counterparts toward young adulthood whereas male left-handers showed reduced performance speed for that age group.

**Figure 5.**
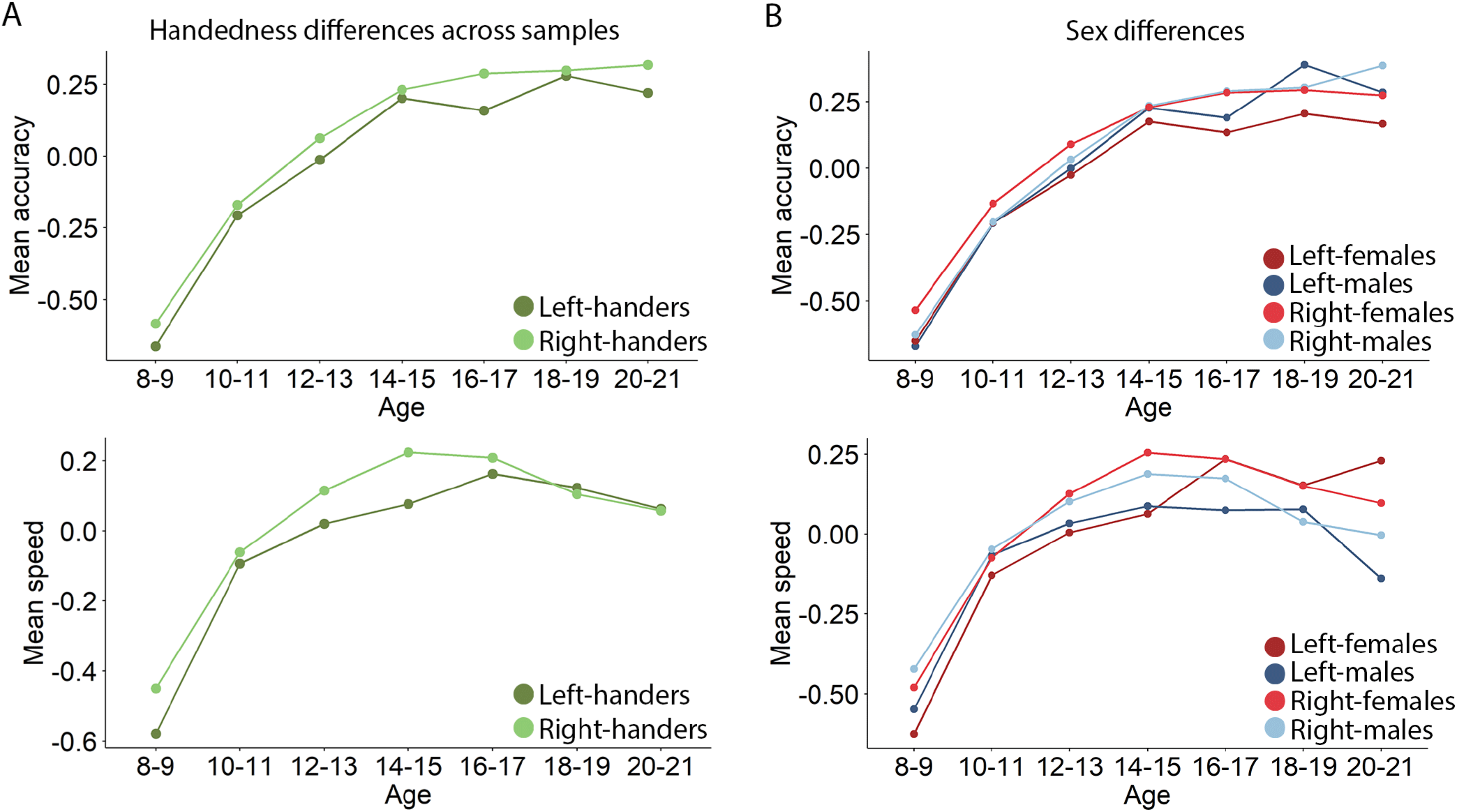
Age trajectory for accuracy (top panels) and speed (bottom panels) of cognitive performance associated with handedness: A. Comparing the two handedness groups across the age range of 8 to 21 years. B. The groups separated by sex.

Regarding the age trajectories of WIV, in Roalf et al (2014), who examined the same data, WIV showed a U-shaped relation with age. Children’s abilities develop unevenly and at different rates, reflected in high initial WIV that decreases toward early adolescence as abilities even out. Toward late adolescence and early adulthood, as specialization occurs, WIV increases again. This increase is more pronounced for speed than for accuracy. Adding age group to the MMRM model showed a strong effect of age (F=166.2, df=6,9120, p<0.001, partial η^2^=0.0986), but no interactions involving age group and handedness. As seen in Figure 6, WIV was higher for left-handers especially for speed in the early age groups and for accuracy in later age groups, and these effects were similar in males and females.

**Figure 6.**
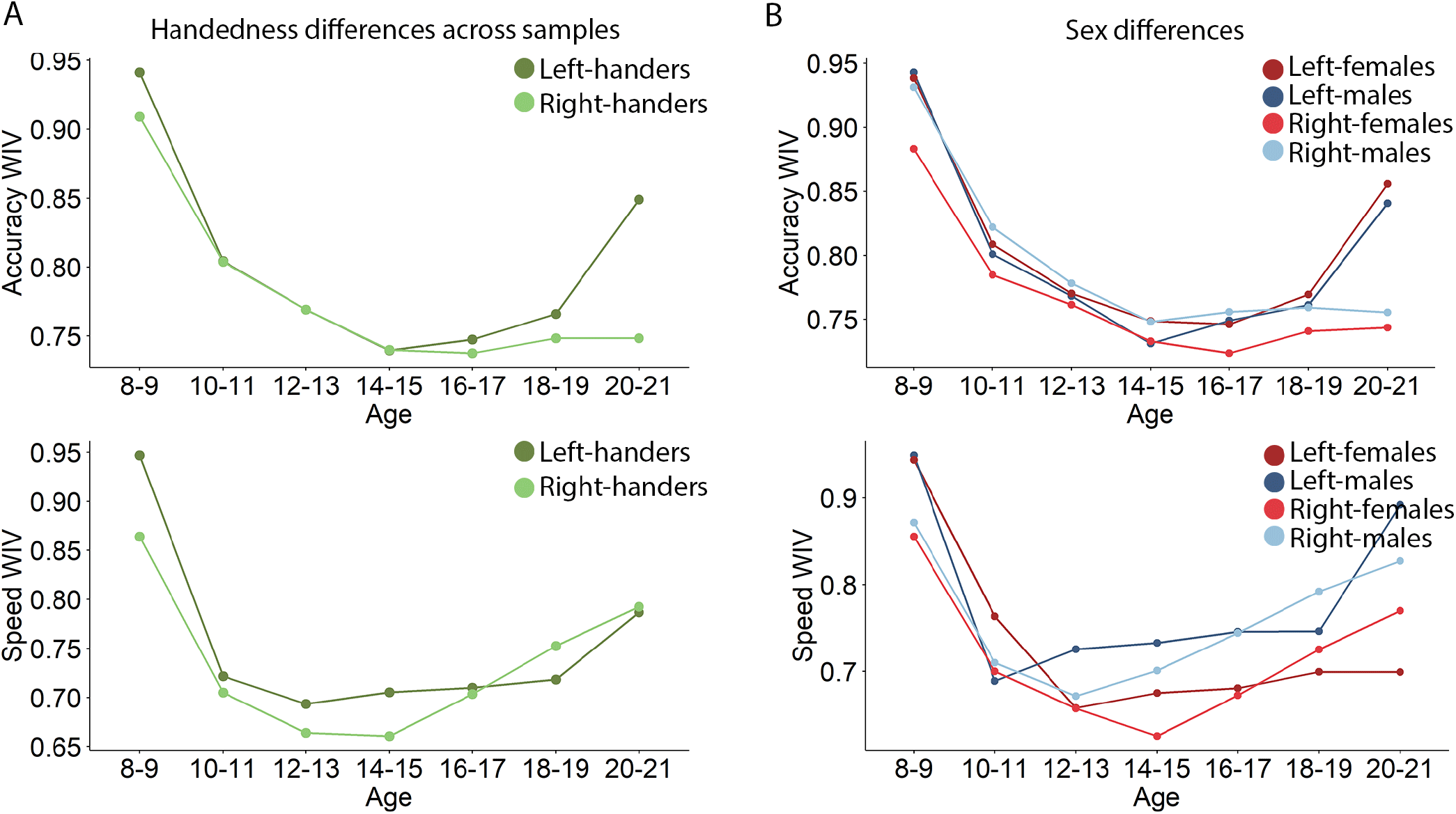
Within-individual variability (WIV) for the handedness groups by age group: A. Comparing the two groups in overall WIV. B. The groups separated by sex.

## Discussion

The environment is symmetric, with an equal opportunity for stimuli requiring responses to occur on the left and on the right. Indeed, most living organisms are symmetric (order Bilateria). However, some species have evolved to sacrifice symmetry in favor of asymmetric organization. These species include hominoids, where the overwhelming majority show right-sided preference reflected in nearly 90% right-handedness in contemporary humans. The adaptive advantage of this sacrifice of symmetry is increasingly understood as allowing brain hemispheric specialization of function. In right-handers, the left hemisphere controls the writing hand and corresponding language functions, while the right hemisphere regulates spatial processes (Mountcasle 1962). This specialization is reflected in behavioral manifestations of brain dysfunction affecting cognitive performance and supported by hemispheric asymmetries in brain structure, neuronal activation, and connectivity (Kurth et al., 2024). Understanding these advantages leaves us wondering about the persistent existence of a minority of non-right handers - both left handers and ambidexters. The perplexity heightens because there is evidence that left-handedness is associated with increased psychopathology and impaired cognitive performance (Packheiser et al., 2025; Abbondanza et al., 2023; Grimshaw and Wilson, 2013). Understanding the behavioral characteristics of left-handers compared to right-handers requires samples in which both groups are represented and assessed with the same clinical and neurocognitive measures. Such samples were not attainable mostly because left-handers have been typically excluded from behavioral studies (Bailey et al., 2020; Cano et al., 2025; Huang et al., 2025; Sriutaisuk and Franz, 2025; Willems et al., 2014). Our study of a community sample of over 9,000 youth ages 8 to 21, unselected for handedness, psychopathology, or cognitive functioning, thus provides a unique opportunity to gain insight into the behavioral profiles of the two handedness groups, examine sex differences, and establish the developmental trajectories of these profiles.

As hypothesized from prior studies, we found that left-handedness was indeed associated with increased psychopathology in all four domains, including mood-anxiety, phobias, externalizing behavior, and psychosis. While the handedness x psychopathology domain interaction was not significant, it is noteworthy that the increased severity for left-handers was significant for externalizing behavior and psychosis, but not for the other domains. This increased psychopathology was evident across the age range of 8 to 21 years but accelerated in early adolescence toward adulthood. There were notable sex differences in the severity of symptoms, females had more severe anxiety-depression and phobia symptoms while males had more severe externalizing and psychosis symptoms. These sex differences were not modulated by handedness.

The association of handedness with cognitive performance was more nuanced. While left-handers performed more poorly than right-handers in most domains, as predicted, they showed equal performance on sustained attention and spatial processing and outperformed right-handers in motor speed. Comparing accuracy with speed of performance revealed an interaction of handedness x test x accuracy vs. speed (p<0.001), indicating that the effects of handedness were stronger for accuracy on some tests and for speed on others. Specifically, left-handers showed deficits for speed and not accuracy for the verbal memory and emotion identification tests, while their deficit was specific for accuracy and not speed for the nonverbal reasoning test. Notably, left-handers showed deficits either on accuracy or on speed on all social cognition tests. This pattern of results would suggest that left-handers might have difficulties with social skills. While there are linguistic clues for such a phenomenon (French “gauche”, German “Linkisch”), and some evidence that atypical sensorimotor biases are associated with social skills (Donati 2024), there is sparse empirical evidence for social skills deficits in left-handers (Johnston et al. 2009) and further research examining this issue is needed. Sex modulated these associations, as demonstrated by the handedness x sex x accuracy vs. speed interaction (p< 0.001). This interaction indicated that the effects of handedness were stronger for accuracy in females than in males while speed was reduced equally. Again, further research is needed to establish the implications of these sex differences.

The hypothesized greater variance in performance distribution in left-handers compared to right-handers was confirmed (p=0.029). However, the handedness groups distributions did not differ in kurtosis, skew, or bimodality (Hartigan dip test). These variance comparison results indicate that the higher variance in left-handers is unlikely caused by an underlying bimodality, as would have been suggested by Satz’s (1972) hypothesis. Rather, the wider variance in left-handers is reflected evenly throughout the distribution of cognitive abilities.

Hypothesis 4 of handedness differences in WIV was strongly supported, with higher WIV for left-handers (p< 0.001). Thus, it appears that left-handers are more likely to be “neurocognitive specialists” rather than “neurocognitive generalists”. There was also a handedness x accuracy vs. speed interaction (p=0.029), indicating that the effects of handedness on WIV were more pronounced on speed than accuracy. This interaction suggests that left-handers are particularly likely to excel in speed in areas where they are performing their best or show speed deficits in areas where they are performing more poorly than right-handers.

Charting the developmental trajectories of the handedness differences as modulated by sex, we found that for the clinical symptoms, adding Age group and Sex as grouping factors to the MMRM showed significant effects of handedness, left-handedness associated with more severe psychopathology (p=0.002), age bin effect (p< 0.001), age associated with increased severity in all symptom domains, but no higher-order interactions. As in previous studies and consistent with the literature, mood and anxiety symptoms, as well as specific phobias, were more prominent in females than males across the age range. They were more prominent in left-handers in both sexes, especially in late adolescence. Again, as expected, externalizing symptoms were more prominent in males than in females across the age ranges, but again both male and female left-handers showed greater severity, especially during adolescence (although severity dropped in left-handed females toward early adulthood). For psychosis, right-handers showed the expectedly higher severity in males across the age range but especially toward young adulthood. Left-handers, both male and female, show increased psychosis symptoms especially during adolescence, with further increase in males toward adulthood. These results indicate that handedness is associated with greater psychopathology throughout the lifespan and in both males and females.

Examining the developmental trajectories of cognitive performance and sex differences in the two handedness groups showed significant effects of age group, older age associated with improved performance efficiency (p< 0.001), and a handedness x age group x accuracy vs. speed interaction (p< 0.001). Both handedness groups showed the expected improvement with age for both accuracy and speed, but left-handers performed more poorly for speed, especially at the earlier age bins. While the 4-way interaction of handedness x sex x age group x accuracy vs. speed was only marginal (p=0.086), it appears that accuracy and speed showed somewhat different trajectories for left-handed and right-handed males and females.

For accuracy, while female left-handers showed lower performance across the age bins, male left-handers outperformed their right-handed counterparts by young adulthood. For speed, female left-handers caught up with their right-handed counterparts toward young adulthood whereas male left-handers showed reduced performance speed for that age group. These more subtle effects require replication and further study.

While WIV was significantly higher in left-handers than in right-handers, the developmental trajectories of WIV were largely similar in the two handedness groups. As in Roalf et al. (Roalf et al., 2014a), who examined the same data, WIV showed a U-shaped relation with age. Children’s abilities develop unevenly and at different rates, reflected in high initial WIV that decreases toward early adolescence as abilities even out. Toward late adolescence and early adulthood, as specialization occurs, WIV increases again. This increase is more pronounced for speed than for accuracy. Indeed, age bin showed a strong effect on WIV (p< 0.001), but no interactions involving age bin and handedness emerged. WIV was higher for left-handers starting in childhood for both accuracy and speed, and these effects were similar in males and females.

Together, our results indicate that left-handedness is characterized by a specific profile of clinical symptoms and neurocognitive functioning. While greater psychopathology was observed in left-handers compared to right-handers across domains, development, and sexes, handedness differences in neurocognitive performance were more nuanced. The hypothesis of greater variance of performance in left-handers was supported, although we found no evidence for greater bimodality. Left-handers performed more poorly on most, but not all domains, and these differences were notable for accuracy in some domains and for speed in others. These differences were also modulated to some extent by sex, as the effects of handedness were stronger for accuracy in females than in males but about equal for speed in the two sexes. The developmental trajectories of cognitive performance measures were notable for greater speed deficits in left-handers at the earlier age groups. The hypothesis of higher WIV in left-handers was strongly supported. The developmental trajectories of these handedness effects were similar in males and females across the age groups.

Current thinking about the ontology of handedness from a genetic perspective (Ocklenburg et al., 2025) is that it arises neither from a monogenetic trait (Annett, 1964), nor solely the environment, but most likely from the contribution of multiple genetic and environmental factors (Corballis, 2010; Ocklenburg et al., 2025). In a study of 54,270 siblings and twins, Medland et al. estimate a 24.64% contribution of genes to handedness with a remainder from non-shared environmental influences (Medland et al., 2009). Consistent with this estimate, Ocklenburg (Ocklenburg et al., 2025) concluded in a recent data analysis that “Handedness heritability is consistently estimated to be 25%.” In evaluating the significance of handedness-related traits, we can examine two complementary principles: the way cognitive and neural features are organized (“packaging”) and how this organization manifests in observable behavior (“performance”), with differences in packaging ultimately shaping differences in performance(Sha et al., 2021).

What is the advantage of left-handedness? Why is there a genetically and environmentally maintained minority for which the other side is more dominant, i.e., a significant minority showing the reverse or a lack of the prevalent asymmetry. There is the obvious advantage of the right-hander who can skillfully attack with the dominant hand while using the other to shield the heart. However, when facing an adversary there could be an advantage to having a minority with exceptional skills on an unexpected side. For example, the tribe of Benjamin had a special unit of left-handed archers sufficient to defeat in battle an army consisting of all other Israel’s tribes (Judges 20:16), and one left-handed Benjamite, Ehud (Judges 3:12-30) took advantage of hiding a dagger on his right thigh to assassinate Eglon, the king of Moab, who had oppressed Israel for 18 years. Notably, left-handers outperformed right-handers on attention, spatial processing, and motor tasks, which require interhemispheric communication. This aspect is consistent with the finding that in left-handers with left hemispheric language, which are likely the majority (Knecht et al., 2000, Szaflarski et al., 2002), even the mundane act of writing requires the left hemisphere literally to “dictate” letter and digit sequences to the right hemisphere (Gur et al. 1984). Because laterality varies along a continuum rather than being an all-or-none trait, the underlying mechanism can be understood as maintaining a functional balance: avoiding excessive symmetry in neural organization while preserving the benefits of asymmetric specialization that allows tasks to be efficiently performed by one side or both (Corballis, 2010; Sha et al., 2021).

Our results indicate that some behavioral characteristics of left-handers could confer additional advantages, specifically a wider range of abilities, better speed of attention and motor speed, and greater neurocognitive specialization. How these advantages translate to real-world adaptation can be a topic worthy of further pursuit, but it requires knowledge about the handedness of individuals and this information is absent in most populations studied.

Worse still, when available, handedness is used to exclude left-handers. In fields where handedness is carefully documented, such as some sports, there is indeed evidence that our findings on the strengths of left-handers are translated to real-world achievement. For example, as ascertained on February 8, 2025, of the 292 members of the Baseball Hall of Fame (excluding non-player Managers), 105 (36%) are left-handed, even though left-handers, while potentially having some advantages (see Harris, 2016), are excluded from four positions (catcher, second base, third base, and shortstop). This ratio far exceeds the expected 10-12% (X^2^=159.066, df=1, p<0.001). Changing attitudes toward left-handers could also play a role.

For example, only 1 of the first 30 US presidents was left-handed, but 7 out of the last 16 were (X^2^=11.89, df=1, p<0.001). Unfortunately, data on handedness is absent or sparse for most professions and further studies in which left-handers are included are necessary to gauge the effects of the behavioral differences we report here on functioning of left-handers in society and their contribution to civilization.

Our findings should be considered in the context of several limitations of the study.

Handedness was determined by self-classification and although this method is highly consistent with more detailed assessments(Bryden 1977, Marchant & McGrew 1998, Da Silva et al., 2012, Gur et al., 1977), it provides no information on the degree of handedness and its relation to other laterality measures such as foot and eye dominance, which are relevant both to psychopathology and to performance (Gur 1978). Further studies are needed to assess the effects of these factors on clinical manifestations and neurocognition. The age range of our study does not allow assessment of developmental trajectories of handedness at younger ages, nor does it allow evaluating effects of handedness on the elderly and aging brain functioning. The cross-sectional design does not permit charting developmental trajectories for individuals and constrains the interpretation of age-related effects. We are also limited to behavioral data, and mechanistic hypotheses would require information on brain structure and function, such as can be obtained with neuroimaging (Sha et al 2021). Multimodal neuroimaging data are available on a subsample of the PNC (Satterthwaite et al., 2014) and we will pursue handedness differences on pertinent neuroimaging parameters in the future.

Notwithstanding these limitations, our study identifies unique profiles of clinical features and neurocognitive performance characterizing left-handers compared to their right-handed counterparts. These unique features may help understand the persistent existence of this significant minority and its potential contribution to civilization. But perhaps more important than any specific finding, our study should serve as a call to brain behavior investigators to incorporate left-handed individuals into their research. We have gone through an epoch when males were almost exclusively studied in behavioral research, this epoch is fortunately over to the benefit of both sexes (Gur and Gur, 2017). Time to end the exclusion of “lefties” to the benefit of all.

## Materials and Methods

Recruitment and clinical, neurocognitive, and neuroimaging procedures for data acquisition, processing, and analysis in the Philadelphia Neurodevelopmental Cohort (PNC) were published previously (Calkins et al., 2015, Gur et al., 2012, 2014, Satterthwaite et al., 2014). The PNC is a genotyped sample recruited from the Children’s Hospital of Philadelphia (CHOP) pediatric network and not from psychiatric services. Enrollment criteria included: stable health; proficiency in English; physical and cognitive ability to complete interview and neurocognitive assessment.

### Clinical Assessment

A structured interview (GOASSESS), incorporating the Kiddie Schedule for Affective Disorders and Schizophrenia (K-SAD), was administered (Calkins et al., 2015). The presence, frequency, and duration of distress or impairment caused by symptoms across psychopathology domains was measured, with algorithms providing DSM-IV diagnosis.

Exploratory item-factor analysis extracting 4 factors (oblimin rotation) was performed on 112 clinical symptom items from the GOASSESS. This analysis produced a clean theory-consistent solution (Anxious-Misery, Fear, Externalizing, Psychosis) that facilitated the building of two confirmatory models from which scores were obtained: a correlated-traits model (4 correlated scores) and a bifactor model (4 orthogonal scores plus an overall psychopathology score, also orthogonal) (see Shanmugan et al., 2016). The former (correlated traits) scores were used here because they are better determined (Grice, 2001) than the bifactor scores, and their collinearity is not a problem because they are treated as correlated dependent variables in a repeated measures mixed model - in contrast to, for example, using the clinical variables simultaneously as independent variables, in which case the bifactor model might have been more appropriate. The PNC is aged 8 to 21 years, is a racially, ethnically, and economically diverse population ascertained from nonpsychiatric pediatric services. Data were acquired from November 5, 2009, through December 30, 2011, concomitantly applying standard protocols. Procedures were approved by the institutional review boards of the University of Pennsylvania and Children’s Hospital of Philadelphia, Philadelphia, Pennsylvania.

For participants under the age of 18 written informed consent was obtained from legal guardians; children provided written assent. This study followed the Strengthening the Reporting of Observational Studies in Epidemiology (STROBE) reporting guideline.

### Neurocognitive Assessment

The Penn Computerized Neurocognitive Battery provided measures of accuracy and response time for executive function, episodic memory, complex cognition, social cognition, and psychomotor speed (Gur et al., 2001, 2010, 2012, Moore et al., 2015). To examine efficiency, accuracy and median response time were transformed to their standard equivalents (z scores), with response time z scores multiplied by −1 (so that higher numbers indicate faster response times and thus better performance). The mean of the accuracy and speed z scores was calculated for efficiency.

## Data Availability

The primary data used in this study are available on DBGaP (https://dbgap.ncbi.nlm.nih.gov/beta/study/phs000607.v3.p2/#study). Other publicly available data sources and applications are cited in the Methods section.

## Code availability

This study used openly available software and codes, specifically R. All R code used for this project is available at https://osf.io/uy3ea.

